# IMPACT OF COVID-19 PANDEMIC LED LOCKDOWN ON THE LIFESTYLE OF ADOLESCENTS AND YOUNG ADULTS

**DOI:** 10.1101/2020.08.22.20180000

**Authors:** Shubhajeet Roy, Sunita Tiwari, Shweta Kanchan, Prashant Bajpai

## Abstract

**OBJECTIVE:** This study was designed to study affect of COVID 19 pandemic on lifestyle of young adults and adolescent.

**METHOD:** Online survey, was conducted in about 1000 respondents in the age group of 13–25 years.

**RESULTS:** Questionnaire based survey showed mean sleeping duration changing from 6.85hours to 8.17hours, average screen time becoming 5.12hours from 3.5hours, 51.9% subjects experiencing increased stress levels, 76.4% subjects experiencing increased food intake and 38.6% subjects had decreased levels of physical activity as per self-monitoring.

**CONCLUSION:** These changes might have long lasting effect on their physical, mental and social health and need counteractive measures to help young people lead a healthy lifestyle during the epidemic and beyond.

## INTRODUCTION

COVID-19 pandemic began from Wuhan, China and spread like wildfire across the globe. Total lockdown consequent to it taken for preventing spread of COVID 19 resulted in changes in daily life schedule, daily physical activity level, changes in daily life schedule, daily physical activity level, mental activity level, food intake level, sleep duration; restricted physical activity, unlimited screen time and circadian rhythm disturbance coupled with minimal exposure to sunlight—the primary determinant of our circadian rhythm^[1]^and erratic food patterns, have serious repercussions on our health and wellbeing, Even after lockdown is lifted, life will not remain the same as seen in studies^[2]^. The present study is an attempt to find out the effect of this pandemic led lockdown on the lifestyle of younger population.

## MATERIAL AND METHODS

The study was a questionnaire-based survey, created using Google Forms, which was circulated by the investigators via various social media apps like – WhatsApp, Facebook, LinkedIn and Instagram. Ethical clearance was obtained before the study. The questionnaire, was based on the Fantastic Lifestyle Questionnaire^[3]^, which was served as an online link. The snowball sampling was used to circulate it widely across India. The online link opened with the question of Informed Consent. The questionnaire included questions on sociodemographic parameters followed by questions related to sleep, screen time, stress level, food intake and physical activity levels. Based on the responses, they were statistically analysed using descriptive analysis tests, to find the impact of this lockdown on the lifestyle of adolescents and young adults.

### INCLUSION CRITERIA

Indian subjects literate enough to understand questionnaire and in the age group 13years to 25years included in the study.

### EXCLUSION CRITERIA

Subjects suffering from any chronic disease like diabetes. Hypertension, Asthma, mental illness and other systemic diseases, were not included in this study.

## RESULT

A total of 1065 subjects were enrolled and they gave their consent to participate in the survey.

The mean ± SD of age was reported as 19.918 ± 3.5388 years. 19% subjects were from rural background, whereas 81% were from urban background. 3.098% subjects were present in junior classes, i.e. below class Xth, 8.92% were Xth pass, 13.521% were XIIth pass, 65.44% were undergraduates, 7.136% were postgraduates and 1.784% had other educational qualifications.

### COMPARATIVE STUDY OF AVERAGE DURATION OF SLEEP BEFORE AND AFTER LOCKDOWN

**TABLE 1:** This table illustrates the changes in sleep duration before lockdown and during lockdown. Sleep duration has been classified broadly into 4 categories and the number and percentage of subjects in both the categories has been shown below. (ORIGINAL)

| AVERAGE NO. OF HOURS OF SLEEP | BEFORE LOCKDOWN |  | DURING LOCKDOWN |  |
| --- | --- | --- | --- | --- |
|  | Number of subjects (n= 1065) | % of subjects | Number of subjects (n= 1065) | % of subjects |
| 4-6 Hours/day | 241 | 22.6% | 83 | 7.8% |
| 6-8 Hours/day | 672 | 63.1% | 420 | 39.4 % |
| 8-10 Hours/day | 140 | 13.1% | 463 | 43.5% |
| >10 Hours/day | 12 | 1.1% | 99 | 9.3% |

Average sleep duration changed from 6.858 hours before lockdown to 8.179 hours during lockdown

### AVERAGE SCREEN TIME DURING LOCKDOWN

The average screen time is increased during lockdown, with average screen time being 5.12985 hours, which is very high when seen in Indian scenarios. Pre lockdown data of Screen time in India is an average of 3.5hours.^[4]^

### IMPACT ON STRESS LEVEL

**FIG 1:**
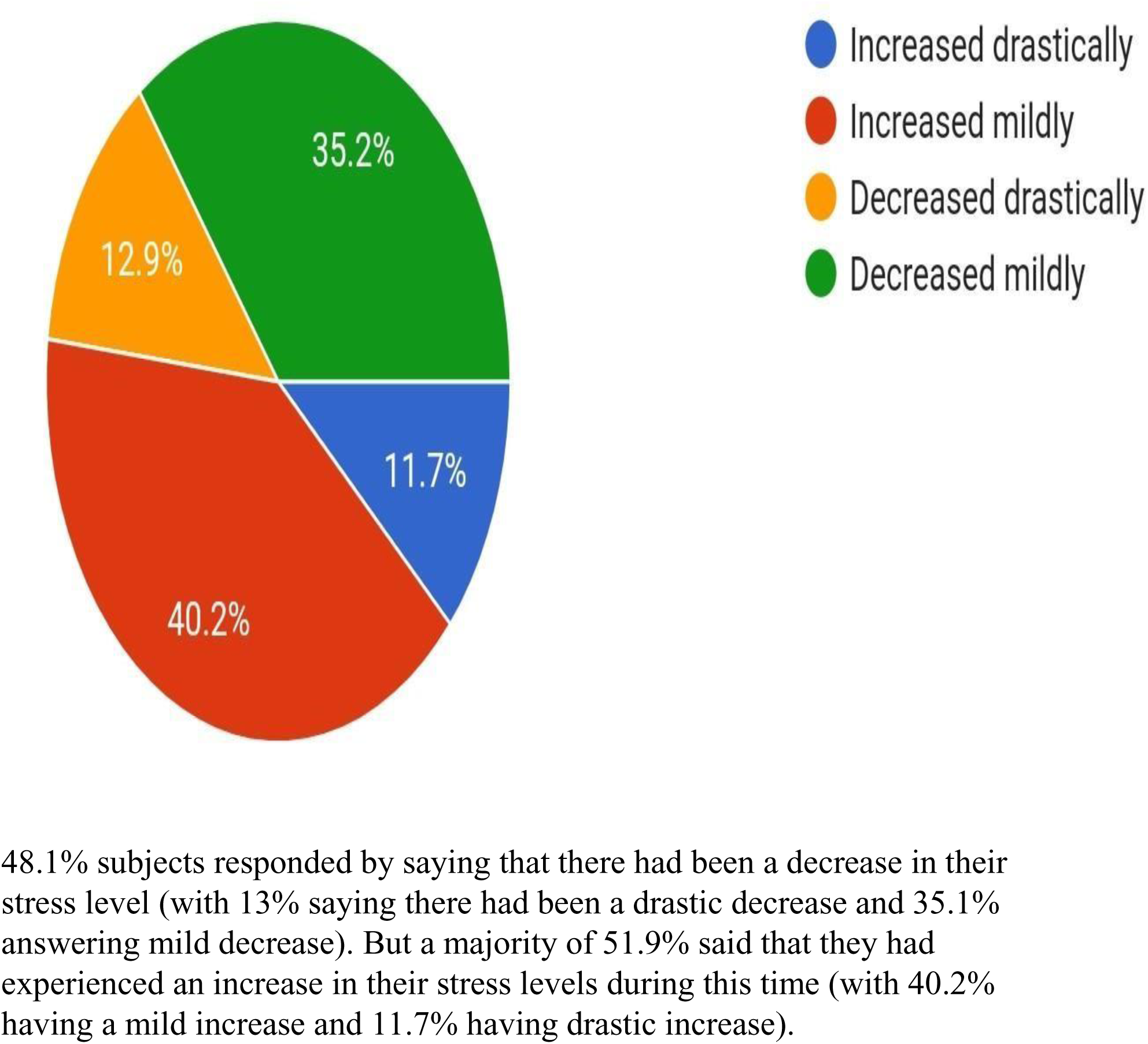
This pie chart shows the changes in stress level of subjects as experienced by them, compared to pre-lockdown conditions. (ORIGINAL)

### IMPACT ON QUANTITY OF FOOD INTAKE

**FIG 2:**
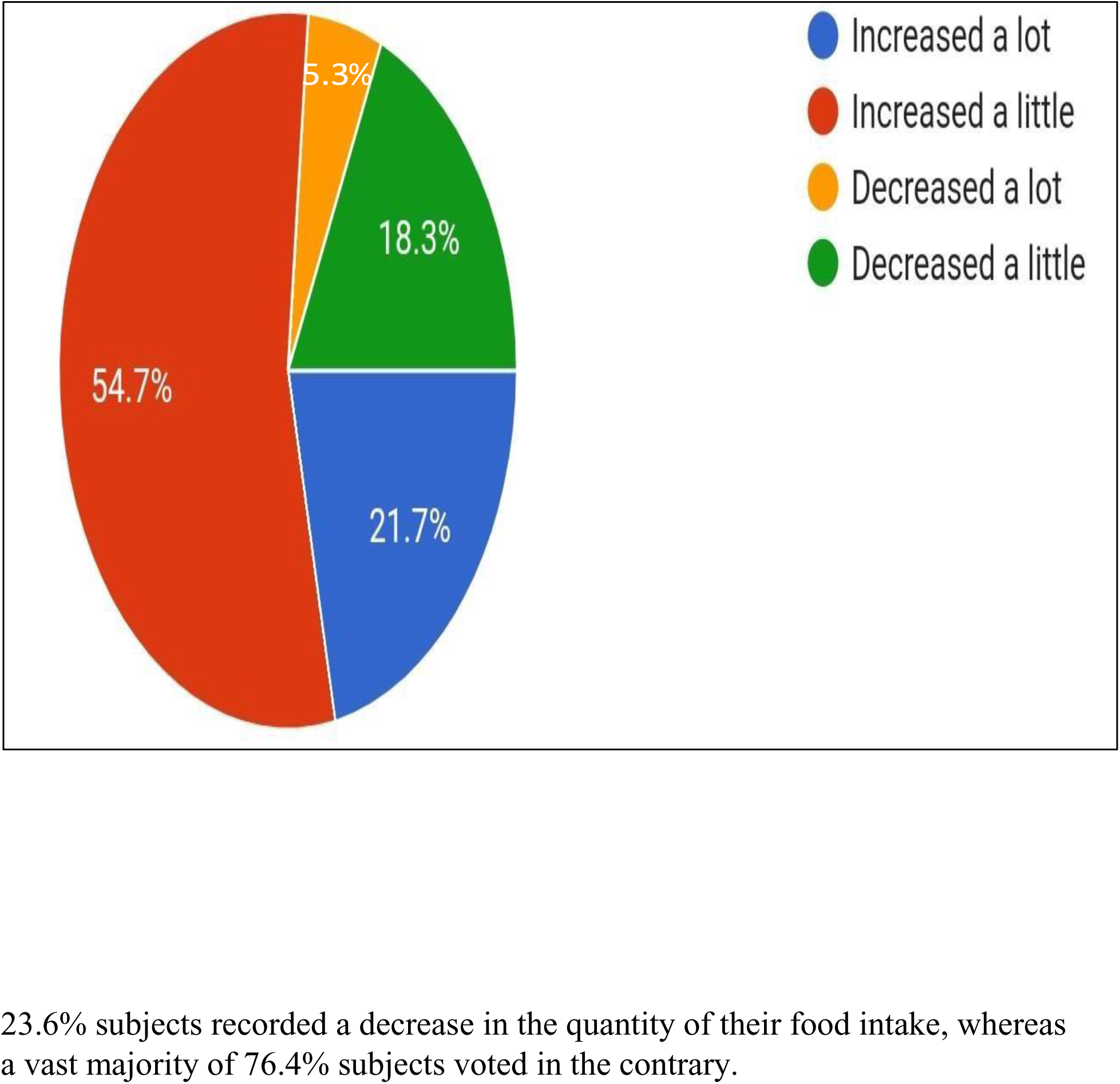
This pie chart shows the changes in food intake of subjects, compared to pre-lockdown conditions. (ORIGINAL)

### LEVEL OF PHYSICAL ACTIVITY DURING LOCKDOWN

**TABLE 2:**
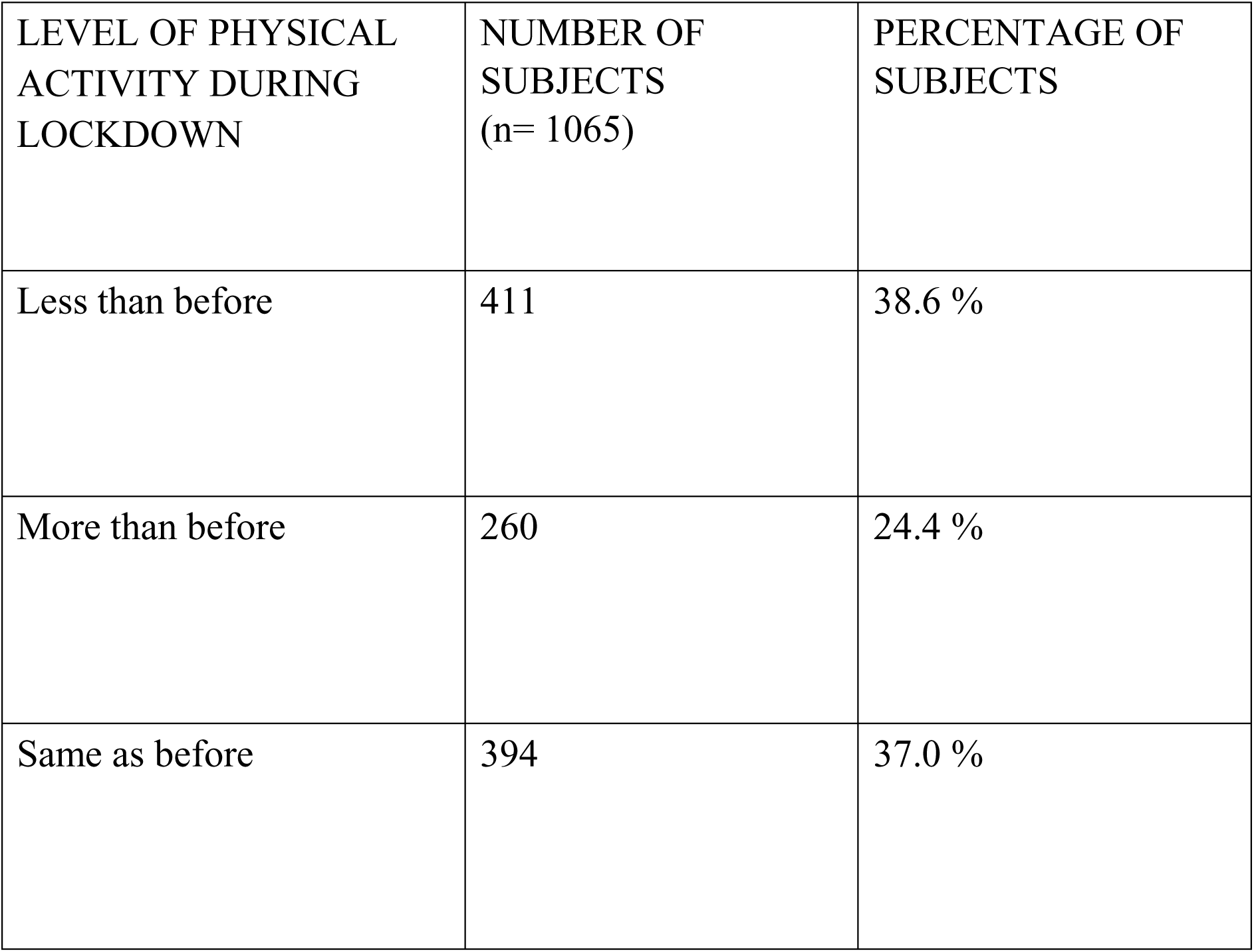
This table shows the response of our subjects, with respect to the level of their physical activity, as compared to pre lockdown conditions. (ORIGINAL)

| LEVEL OF PHYSICAL ACTIVITY DURING LOCKDOWN | NUMBER OF SUBJECTS (n= 1065) | PERCENTAGE OF SUBJECTS |
| --- | --- | --- |
| Less than before | 411 | 38.6 % |
| More than before | 260 | 24.4 % |
| Same as before | 394 | 37.0 % |

The above table clearly shows that the level of physical activity has decreased for greater number of subjects (38.6%) or it has remained the same, as seen in 37% subjects.

We also asked our subjects whether they have tried anything creative or have learnt anything new during this time, to which 79.3% replied in the affirmative.

## DISCUSSION

Results of the present study show a clear increment in the sleeping duration in majority of the subjects. This might be simply due to the fact that there is increased availability of leisure time. Although the quality of this extra duration of sleep remains to be seen.

The increase in screen time as observed in the study, which might be due to the rapid surge in online classes by educational institutes, in an effort to cope up with the back-log of classes due to this lockdown. Moreover, increased free time for the youth has led to the increased time that they are spending over social media apps and also increased rate of binge watching. The lockdown has also increased smartphone usage with weekly/daily time spent per user on smartphone increasing 13%/12% after 3 weeks of COVID-19 outbreak. Daily time spent per user on video streaming platforms (VOD) increased in parallel to smartphone usage. New apps and websites also clocked growth via smartphone. ^[5]^

The increase in stress level, which happened with the majority can be associated to many cause like: first and foremost threat of infection from the dreadful virus and the overload of negative news from various kinds of media sources which has created a negative ambience all around; also there is the threat of losing their near and dear ones. Secondly, unemployment and loss of family income owing to the lockdown can also be a major cause of worry and stress. Also, for the educated youth, the big question mark over their future prospects and opportunities and lack of infrastructural advantages can also be a major stressor. Most reviewed studies reported negative psychological effects including posttraumatic stress symptoms, confusion, and anger. Stressors included infection fears, frustration, boredom, inadequate supplies, inadequate information, financial loss, and stigma. Some researchers have suggested long-lasting effects. ^[6]^ Our results were similar to those found by Robert Stanton, Quyen G. To, SamanKhalesi et al. ^[7]^ A deeper analysis, also leads to revelation of the fact that during this time due to the homeostatic and neuroendocrine disbalance, due to disbalance in circadian rhythms and stress, there might be increased release of the hormone Ghrelin. ^[8]^ And Ghrelin induces hunger greatly. ^[9]^ The matter of concern is the fact that the coupled increase in food intake and decrease in physical activity levels can have debilitating effects on the overall health of a being—starting from weight gain and obesity and finally leading to severe complications like Diabetes, Cardiovascular Diseases and other serious conditions. ^[10]^ Recognizing the adverse collateral effects of the COVID-19 pandemic lockdown is critical in avoiding depreciation of weight control efforts among youths afflicted with excess adiposity. Depending on duration, these untoward lockdown effects may have a lasting impact on a child’s or adolescent’s adult adiposity level. ^[11]^ Increased screen-time coupled with increasing stress levels and decreasing physical activity levels, can have their due repercussions on their ophthalmic health like narrowed retinal micro vascular structure ^[12]^ as well as leading to an array of other psychological, cognitive and physical disorders like obesity, sleep problems, depression and anxiety, to name a few. ^[13]^The study could be made more elaborative with a larger sample size and a longer follow up for residual side effects of lock down.

## CONCLUSION

The above study showed changes in the various lifestyle indices in younger population owing to the COVID 19 pandemic led lockdown. The increased screen time and habits like binge watching could be countered by an encouragement of co-curricular activities in the adolescent and young adults. Indoor physical activity and outdoor activity with due precautions should be encouraged by parents and educational institutes as a part of their online education. The course of the pandemic is uncertain, and may last long. Young generation should maintain a fixed sleep wake schedule, healthy eating habits and some degree of exercise regime while following the safety norms to prevent the spread of contagion, only then can the youth emerge victorious and expand their horizons even under the restrictions imposed by the corona virus.

## Data Availability

ALL REFERENCE DATA WAS COLLECTED USING GOOGLE SCHOLAR AND PUBMED.
ALL STATISTICAL DATA WERE COLLECTED BY OURSELVES.

## ACKNOWLEDGEMENT

In the conduction of this study, we were helped in a number of stages by few people we would like to mention. In the data collection process, we were helped a lot, by three undergraduate students, namely: Ms Nikita Singh, Mr Siddhant Aggarwal and Ms Devanshi Katiyar.

## Notes

### Competing Interest Statement

The authors have declared no competing interest.

### Funding Statement

NONE

### Author Declarations

INSTITUTIONAL ETHICS COMMITTEE FOR HUMAN RESEARCH, KING GEORGE'S MEDICAL UNIVERSITY, LUCKNOW, INDIA.

